# Assessment of Knowledge, Attitude, and Practice Towards the First Aid Management of Foreign Body Aspiration and Obstruction Among Parents of Children Visited Sphmmc, Addis Ababa, Ethiopia

**DOI:** 10.1101/2024.10.31.24316500

**Authors:** Yohannes Hailu, Sisay Yifru

## Abstract

**Background:** Foreign body aspiration and obstruction are the main cause of accidental death in childhood, and Foreign bodies (FBs) in the aerodigestive tract provide diagnostic and therapeutic problems and are significant sources of morbidity and mortality. Lack of community awareness toward the FBA presentation and its management further aggravate the problem. Despite this, there are very few studies in this area which necessitates further study. Therefore the present study may help as an output to increase parents’ awareness towards immediate first aid intervention, in preventing delayed healthcare access and intervention at a community level in general.

**Objective:** Assessing the knowledge, attitude, and practice towards the first aid management of foreign body aspiration and obstruction among parents of children coming to St. Paul’s Hospital Millennium Medical College.

**Methods:** A descriptive-based cross-sectional study was carried out in the study area using pretested, structured, and self-administered questionnaires. The collected data was analyzed using SPSS version 25. Multiple logistic regression analysis was used to identify factors associated with the Knowledge, attitude, and practice of parents towards first aid management of foreign body aspiration and obstruction.

**Results:** A total of 423 parents were involved in the study. Only 218 (51.5%) of them were knowledgeable. Most of the respondents (76.3%) had a positive attitude towards foreign body that foreign body aspiration needs immediate intervention, and 75 (17.7%) faced a child who aspirated foreign body. Of these, only 53 (70.7%) had provided first aid to the victim. Most of the respondents 40 (75.5%)had scored below 80% of practice towards foreign body aspiration and obstruction. Multiple Logistic Regression Analysis showed that parents who are literates were 3.6 times more knowledgeable than those illiterates. (AOR:3.612, 95% CI: 1.758, 7.420, P*≤*0.05).

**Conclusions:** The level of knowledge and skills for providing first aid for foreign body aspiration, and obstruction in children among parents are low. So education and increasing awareness among them to reduce morbidity and mortality in children suffering from aspiration and obstruction from foreign bodies has to be one of the strategies.

## Background

Inhaling foreign bodies or items into the respiratory tract is known as foreign body aspiration, or FBA. [1] FBA is frequently seen in pediatric populations. [2] Children often use their tongues to investigate new objects when they are between the ages of six months and four years old. [3] A variety of foreign objects have been documented in the literature; toys, candies, gums, batteries, rocks, and magnets are the most commonly aspirated alien objects. [4] In our setup a study showed plastic tips, seed, metallic tips, balloon inflating tips, peanuts, chicken peas, and hijab pins in our setup. (25) Pediatric FBA is a potentially fatal illness that claims many lives each year, particularly in children under the age of two. [5] Foreign body obstruction can result in compromised ventilation and oxygenation, which may cause a considerable amount of morbidity or death. Pulmonary hemorrhage is the second most common cause of mortality, behind hypoxic-ischemic brain damage.[6,7] Neurologic impairment, pulmonary abscess, bronchiectasis, and recurrent pneumonia are Complications following FBA. [8–10] A comprehensive retrospective assessment conducted in the United States in 2016 found that the death rate for pediatric patients with FBA was 2.5 percent. [8] an aspiration of a foreign body is a dramatic event that can have deadly consequences. The most effective preventive strategy to lower the problem’s incidence is education. [25]. According to the National Safety Council, there are 0.43 and 20.4 fatal and nonfatal choking incidents for every 100,000 American children in the general population, respectively. [9]

The prevalence of foreign body aspiration (FBA) in our study setting is unknown. A previous study conducted in our area involving 81 patients revealed that only 18% of cases were reported within 24 hours of the occurrence, while 82% were reported after that time. Reasons for the delayed presentation included delayed transportation, delayed detection, and parents’ or caregivers’ lack of awareness and of them, 93.8% experienced complications, such as pneumothorax, atelectasis, and lung infections.[19].

The prompt identification and appropriate management of FBA have produced in a significant reduction in the morbidity and mortality rates associated with FBA [10–13].

Parents are the primary caregivers for young children and FBA is a potentially fatal yet avoidable disorder in the pediatric age group that calls for greater parent education and awareness. One significant factor putting kids at increased risk for FBA and related problems is parents’ lack of knowledge about risk factors and how to prevent FBA. To date, there are no published reports on awareness among parents about FBA in children in our local setting.

## Methods

### Study setting and period

This study was conducted at St. Paul’s hospital millennium medical college. SPHMMC is established in 1961 G.C. as a medical college and is a tertiary hospital under federal minister of health Ethiopia. With over 2800 staff, it provides clinical and preclinical training and specialized services. The pediatrics department is a core area, providing outpatient and inpatient services and academic activities. The study was conducted from September 1, 2023, to October 30, 2023 GC.

### Study Design

A descriptive-based cross-sectional study was carried out on parents of children visiting St Paul Hospital Millennium Medical College, from September 1, 2023 to October 30, 2023 GC.

### Sampling

Parents of children who visited SPHMMC were included in the study. A convenient sampling technique was used. Parents whose children were severely ill (admitted to ICU or ED(Red)) were excluded from the study.

The sample size was calculated using a single population proportion formula n = (Z α/2)^2^ x P(1-P) /d2, by taking the assumptions as Zα/2 = 1.96 (standard normal value corresponding to 95% level of confidence), p = 0.5 (estimate of prevalence for KAP of foreign body aspiration first aid to be 50%, since there is no similar study conducted in the study area), and d (margin of error) =5%, so the n value is 384 and after adjusting 10% for possible nonresponse rate made the final sample size 423.

### Data Collection Instruments and Techniques

A self-structured and designed questionnaire was used, which was pre-tested before use for the present study. A questionnaire from previous investigations was modified and used [17, 20, 28]. The questionnaire has four parts. It included demographic profile (age, gender of parent, age of youngest child, number of kids, level of education of parents), questions regarding basic knowledge about foreign bodies (knowing about common age groups, foods that may cause FBA (carrot, seeds, walnut, hazelnut, grapes, cucumber, hard nuts), small toys, leaving kids unsupervised, letting the child hold the toy while crying, talking, or laughing), knowledge about the prevention of FBA, and first aid given after foreign body ingestion. The educational status was stratified into literate and illiterate, where illiterate means a person who is unable to read and write. The questionnaire was translated into Amharic as well, and the data collector explained the respondent and recorded the responses.

### Operational Definitions

**Foreign body aspiration**: the act of inhaling or breathing foreign bodies into the air system or aerodigestive system

**Adequate knowledge**: a participant who scored a mean and above for knowledge questions.

**Inadequate knowledge**: a participant who scored below the mean for knowledge questions.

**Positive Attitude**: a participant who answered agree and strongly agree for attitude questions and scored mean or above

**Negative attitude**: a participant who answered disagree and strongly disagree for attitude questions and scored below the mean.

**Good Practice**: a participant who scored 80%and above on practical questions (according to AHA pediatric basic life support and advanced life support 2020)

**Poor practice**: a participant who scored below 80%of practical questions (AHA, PBLS and PALS 2020)

**First aid**: help given to a foreign body aspirated child until full medical treatment is available.

**Critically ill**: those who are admitted to PICU and at ED (red patients)

**Illiteracy** :unable to read and write

### Data Analysis

Parents Knowledge of first aid provision for foreign body aspiration and obstruction was assessed using nine questions. The questions were dichotomized into knowledgeable and not-knowledgeable. A score of the mean value or above was considered knowledgeable, while a score less than the mean value was taken as not-knowledgeable. Knowledge was taken as a dependent variable, and independent variables were the sex and age of participants, educational status, and occupation. The variables were taken from previously studied literature. Parents’ attitude towards first aid management for a choking child was assessed using a four-point Likert scale. Strongly agree-4, agree-3, disagree-2, and strongly disagree-1. A score of 3 or 4 was given. For positive attitude, while 2 and 1 were for negative attitude, and the order of scoring for negative statements was reversed. Then, the score was dichotomized into positive and negative attitudes for each question. The practices of teachers were described and summarized using descriptive statistics.

The data was analyzed using the Statistical Package of Social Sciences (SPSS) version 27.0 for Microsoft Windows. Means, percentages, and ratios were calculated. Univariate and bivariate analyses between dependent and independent variables were performed using binary logistic regression. A multivariate analysis was done to control for the possible confounding variable. Microsoft Excel 2019 was used to make graphs and charts. In this study, all tests with a P-value < 0.05 were considered statistically significant.

The results were presented using charts, tables, and texts.

### Ethical Consideration

Ethical clearance was obtained from the institutional review board of St Paul’s Hospital Millennium Medical College. Parents were informed about the aim, benefits, and possible inconveniences of the study. They were assured the information they gave would be confidential. Before data collection begins, verbal consent is obtained, and participants can withdraw at any time if the need arises.

## Results

### Socio-demographic characteristics

As shown on table 1,in this study, a total of 423 participants were involved, and the response rate was 100%. Most respondents, 225 (53.2%), were female, with a female-to-male ratio of 1.15:1. The average age of the respondents was 32.41 years. Regarding the educational level of the participants, the majority (379; 89.6%) are literate, 44 (10.4%) are illiterate, 43 (10.2%) are health professionals, and 380 (89.4%) are non-health professionals. The majority of participants (308; 72.8%) have more than 3 children, and most of them (201; 47.7%) are in the youngest child age group of 1–5 years.

**TABLE 1.** 1. Socio-demographic characteristics.

1. Socio-demographic characteristics
| **Variable** |  | **Frequency** | **Percentage** |
| --- | --- | --- | --- |
| Gender | 1.Male | 225 | 53.2% |
|  | 2. Female | 198 | 46.8% |
| Age | 1.18-30 | 213 | 50.3% |
|  | 2.31-50 | 171 | 40.4% |
|  | 3.Above 50 | 39 | 9.2% |
| Marital status | 1. Married | 353 | 83.5% |
|  | 2. Single | 48 | 11.3% |
|  | 3. Divorced | 21 | 5% |
|  | 4. Widowed | 1 | 0.2% |
| Religion | 1. Orthodox | 218 | 52% |
|  | 2. Muslim | 103 | 24.3% |
|  | 3. Protestant | 88 | 20.8% |
|  | 4. Catholic | 9 | 2.1% |
|  | 5.Others | 3 | 0.7% |
| Level of Education | 1.literate | 379 | 89.6% |
|  | 2.illiterate | 44 | 10.4% |
| Occupation | 1. Health professional | 43 | 10.2% |
|  | 2. Non-health professional | 378 | 89.4% |
| Number of children | 1.1-3 | 114 | 27% |
|  | 2.>3 | 308 | 72.8% |
| Age of the youngest child | 1.<1 year | 146 | 34.5% |
|  | 2.1-5 years | 201 | 47.5% |
|  | 3.6-10 years | 49 | 11.6% |
|  | 4.>10 years | 27 | 6.4% |

### Knowledge on first aid management of FBA and Obstruction among parents

The mean *±* SD knowledge score was 5.51 (1.02)/9 points, and only 51.5% of parents scored above the mean value of knowledge questions. Forty-eight (11.3%) of participants had a child aspirate a foreign body, and two hundred and thirty (68.8%) heard about foreign body aspiration, and two hundred twenty-seven (53.7%) heard about first aid for foreign body aspiration and obstruction. The majority of parents 276 (65.2%) correctly identified that children aged 1–5 years were at the highest risk of FBA. Most 320 (75.7%) agreed that children should not be given peanuts until they reach 4 years old. On the other hand, half of them (49.2%) disagreed the idea that the absence of choking is an assuring sign that the item is gone, and a majority proportion (68.3%) even more disagreed with the statement, “If the foreign body causes no symptoms, it is okay to delay removal.” Furthermore, 82.7% of parents agreed that talking while chewing may lead to aspiration. Additionally, a great proportion of parents (67.8%) believed that both non-organic and organic items could cause aspiration in children.

**TABLE 2:** Knowledge on FBA and Obstruction among parents.

| Variables | Response | Frequency | Percentage |
| --- | --- | --- | --- |
| Have any of your children aspirated a foreign body? | 1. Yes | 48 | 11.3% |
|  | 2.No | 375 | 88.7% |
| Have you heard about foreign body aspiration? | 1. Yes | 293 | 68.8% |
|  | 2.No | 130 | 30.7% |
| Heard about first aid related to FBA and FBAO | 1. Yes | 227 | 53.7% |
|  | 2.No | 195 | 46.1% |
| Children can aspirate foreign bodies. | 1. Yes | 366 | 86.5% |
|  | 2.No | 58 | 13.5% |
| At what age children are at the highest risk to aspirate foreign bodies? | 1. <1 year | 116 | 27.4% |
|  | 2. 1-5 years | 276 | 65.2% |
|  | 3. 6-10 years | 30 | 7.1% |
|  | 4. >10 years | 1 | 0.2% |
| Children shouldn't be offered peanut seeds until they are 4 years old | 1. Yes | 320 | 75.7% |
|  | 2.No | 103 | 24.3% |
| The absence of choking is an assuring sign that the item has gone away | 1. Yes | 215 | 50.8% |
|  | 2.No | 208 | 49.2% |
| If the foreign body causes no symptoms it is okay to delay the removal | 1. Yes | 134 | 31.7% |
|  | 2.No | 289 | 68.3% |
| Talking while chewing may lead to aspiration | 1. Yes | 350 | 82.7% |
|  | 2.No | 73 | 17.3% |
| X-rays can detect all foreign bodies | 1. Yes | 260 | 61.5% |
|  | 2.No | 163 | 38.5% |
| Which of the following items are children at risk to aspirate? | 1. Organic like nuts, seeds | 119 | 28.1% |
|  | 2. Non-organic like small plastic toys | 15 | 3.5% |
|  | 3. Both | 287 | 67.8% |
| What types of symptoms do you observe from a person facing foreign body aspiration and obstructed? | 1. Unable to stand (falling) | 26 | 6.1% |
|  | 2. Difficulty of producing sounds | 79 | 18.7% |
|  | 3. Inability to cough forcefully | 59 | 13.9% |
|  | 4. Hands clutched to the throat | 36 | 8.5% |
|  | 5. Difficulty of breathing | 134 | 31.7% |
|  | 6. All | 89 | 21% |

### The attitude of parents on first aid management of foreign body aspiration and obstruction

The mean *±* SD attitude score was 13.8 (4.29)/24 points. Of the total participants, the majority (76.7%) of them have a positive attitude towards providing first aid for a child with foreign body aspiration and obstruction. 233 (55.1%) and 123 (29.5%) of participants strongly agreed and agreed that foreign body 140 (33.1%) strongly agreed and agreed that everybody should know about first aid management of foreign body aspiration, and obstruction, respectively. 164 (38.8) participants strongly disagree that choking causes death or a life-threatening condition if not treated. 127 (37%), agreed that it was possible to manage foreign body aspiration at home without taking the victim to the health institution. 92 (21.7%) strongly disagree that they should sweep their fingers blindly into the throat of a foreign body-aspirated victim, and 85 (20.1%) strongly agree with it. The majority of respondents, 125 (29.5%) and 121 (23.9%) disagreed or strongly disagreed with providing first aid without proper knowledge on how to do it.

**TABLE 3:** Attitude on first aid management of FBA and Obstruction among parents.

| Statement | Strongly Disagree | Disagree | Agree | Strongly Agree |
| --- | --- | --- | --- | --- |
| Foreign body aspiration should need immediate management | 46 (10.9%) | 20 (4.7%) | 123 (29.1%) | 233 (55.1%) |
| Everyone should know about first aid management of foreign body aspiration and obstruction | 41 (9.7%) | 39 (9.2%) | 140 (33.1%) | 203 (48%) |
| Foreign body aspiration does not cause death/life-threatening conditions even if not treated | 164 (38.8%) | 157 (37.1%) | 45 (10.6%) | 57 (13.5%) |
| It is possible to manage foreign body aspiration and obstruction at home without taking a victim to the health care setup | 87 (20.6%) | 176 (41.6%) | 127 (30%) | 33 (7.8%) |
| We should sweep our fingers blindly into the throat of foreign body aspirated and obstructed victim & take the victim to health Institution | 92 (21.7%) | 100 (23.6%) | 146 (34.5%) | 85 (20.1%) |
| You must not provide foreign body aspiration and obstruction first aid without knowledge | 101 (23.9%) | 125 (29.6%) | 124 (29.3%) | 73 (17.3%) |
| If first aid is not given to foreign body obstruction within golden time, it may lead to death | 59 (13.9%) | 21 (5.0%) | 173 (40.9%) | 170 (40.2%) |

### Practice of parents on first aid management of foreign body aspiration and obstruction

Of the participants, 75 (17.7%) faced foreign body-aspirated or obstructed children. Among these, 53 (70.6%) had provided first aid. Based on practice for a child chocking with an object not visible, 9 (17%) gave a glass of water, while 5 (9.4%) did a finger sweep to identify and remove the object, and 14 (24.5%) slapped at the back of the neck, 11 (20.8%) did an abdominal thrust, and 14 (26.4%) slapped at the back, and 1 (1.9%) took the child to the hospital. Based on the assessed practical question for which a child obstructed, developed talking and breathing difficulties with a visible and accessible foreign body, 8 (15.1%) gave a glass of water, while 13 (24.5%) did a finger sweep to identify and remove the object, 10 and slapped at the back of the neck, 10 (18.9%) did an abdominal thrust, 11 (20.8%) slapped at the back, and 1 (1.9%) did not know what to do or took to the hospital. Based on the assessed practical question for a child with choking and coughing only 10 (18.9%) gave a glass of water, while 6 (11.3%) did a finger sweep to identify and remove objects, 7 and slapped at the back of the neck, 10 (18.9%) did an abdominal thrust, 18 (34%), slapped at the back, and 2 (3.8%) didn’t know what to do.

Based on the assessed practical question for child became obstructed, became breathless, and became unconscious. 15 (28.3%) did CPR, while 8 (15.1%) did a finger sweep to identify and remove objects, and 5 (9.4%) slapped at the back of the neck, 7 (13.2%) did an abdominal thrust, 8 (15.1%) slapped at the back, and 10 (18.9%) did not know what to do or took to the hospital.

Overall, 75.5% of study participants scored below 80% of practice towards foreign body aspiration and obstruction, and they are considered to have poor practice towards first aid management of foreign body aspiration and obstruction.

**TABLE 4.**
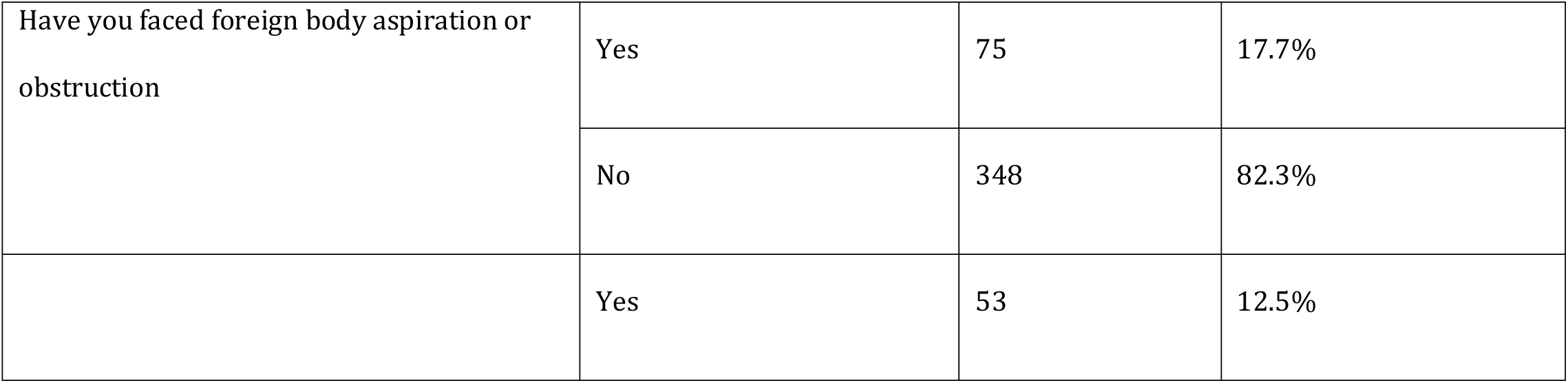
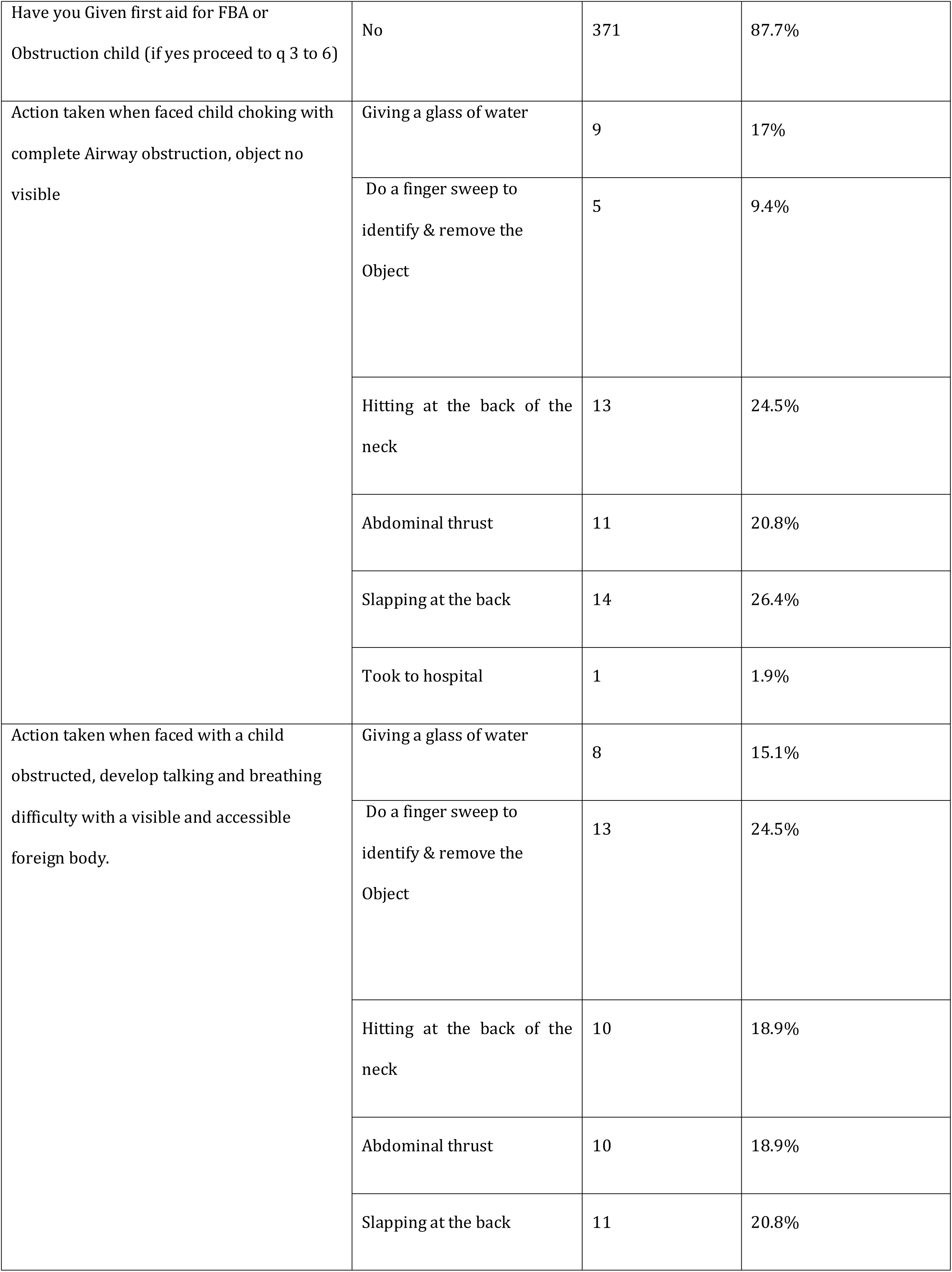
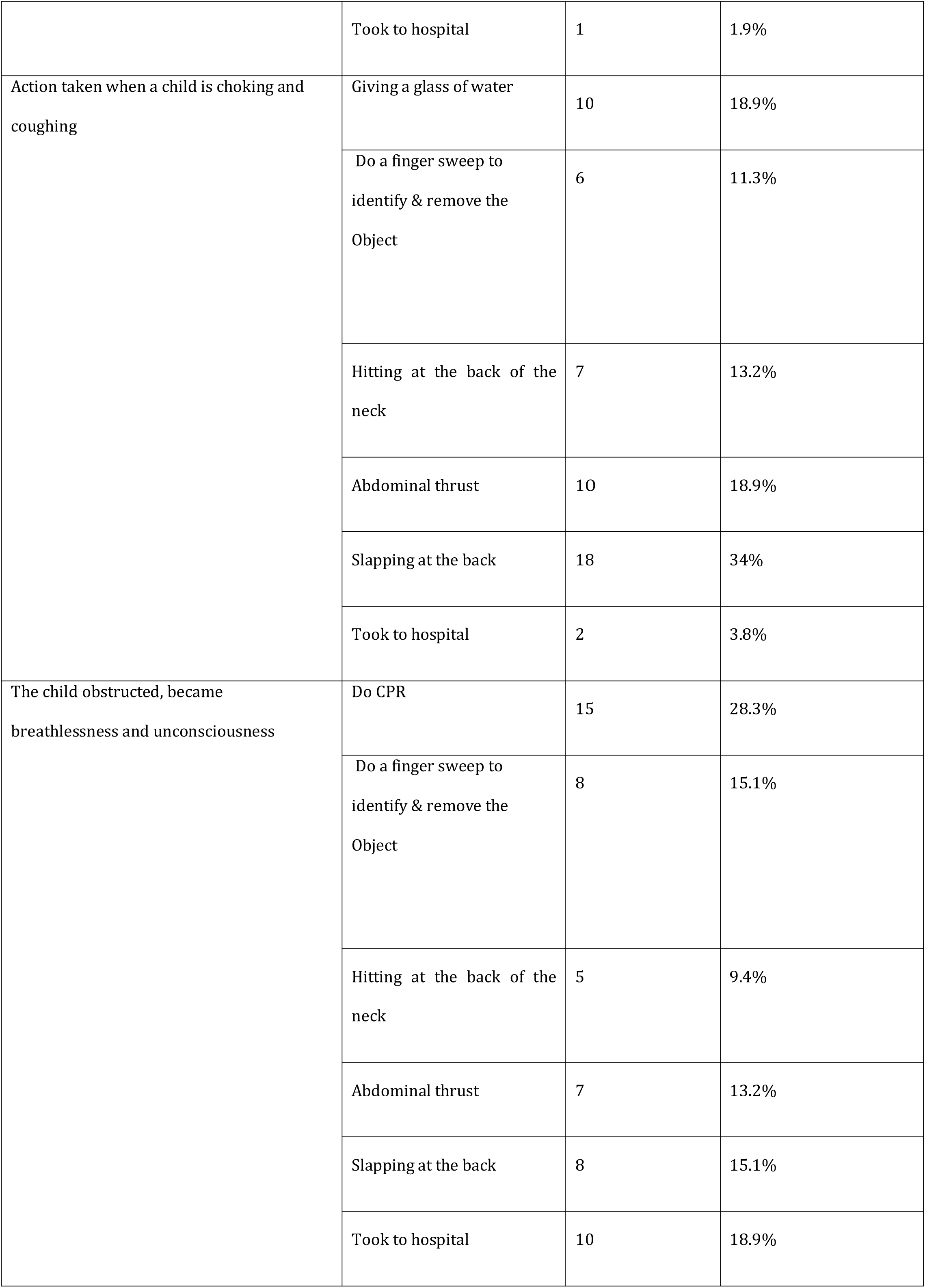
Response of parents practice on FBAO first aid management.

### Factors affecting the knowledge, attitude and practice of parents on first aid management of FBA and Obstruction

Binary and multiple logistic regression analyses were done to analyze factors associated with Knowledge, attitude, and practice in providing first aid management towards foreign body aspiration and obstruction. In the binary logistic regression analysis, sex, age, occupation, and educational status were all associated with knowledge, attitude, and practice of first aid management for a foreign-aspirated child. Parents’ knowledge of first aid management of foreign body aspiration and obstruction was significantly correlated with literacy and health professionalism (AOR:3.612, 95% CI: 1.758, 7.420, P *≤*0.05) and (AOR:3.166, 95% CI: 1.453, 6.263, P*≤*0.05), respectively. Health professionals were significantly associated with parents good practices (AOR: 3.317, 95% CI: 0.834, 12.263, P *≤*0.05). The demographic data of parents has no association with their attitude and practice of FBAO first aid.

**TABLE 5:** Binary and multiple logistic regression analysis of selected factors affecting knowledge on FBA and obstruction.

| Variables |  | Knowledge level |  | Linear regression (95% CI) |  |  |  |
| --- | --- | --- | --- | --- | --- | --- | --- |
|  |  | Adequate | inadequate | COR(p<0.25) | p-value | AOR(P<0.05) | p-value |
|  |  | Freq(%) | Freq(%) |  |  |  |  |
| Sex | Male | 103(24.3) | 95(22.5) | 1 |  | 1 |  |
|  | Female | 117(27.7) | 108(25.5) | 1.010(0.836-1.196) | 0.997 | 1.075(0.721-1.623) | 0.722 |
| Age | <30 years | 104(24.6) | 109(28.8) | 1 |  | 1 |  |
|  | 30-50 years | 109(28.8) | 89(21) | 0.875(0.268-2.851) | 0.824 | 0.591(0.160-2.185) | 0.431 |
|  | >50 years | 7(1.7) | 5(1.18) | 1.400 | 0.566 | 1.335 | 0.682 |
| Education | Illiterate | 11(2.6) | 33 (7.8) | 1 |  | 1 |  |
|  | Literate | 209(49.4) | 170(40) | 3.688(1.810-7.515) | <0.001+ | 3.612(1.758-7.420) | <0.001++ |
| Occupation | Non health professional | 188(44.4) | 192(45.3) | 1 |  | 1 |  |
|  | Health professional | 32(7.6) | 11(2.6) | 2.971(1.455-6.067) | 0.003+ | 3.016(1.453-6.263) | 0.003++ |

++ = significant at $p \leq 0.05$
+= associated at $p \leq 0.25$ , COR-crude odds ratio, AOR-adjusted odds ratio.

**TABLE 6:**
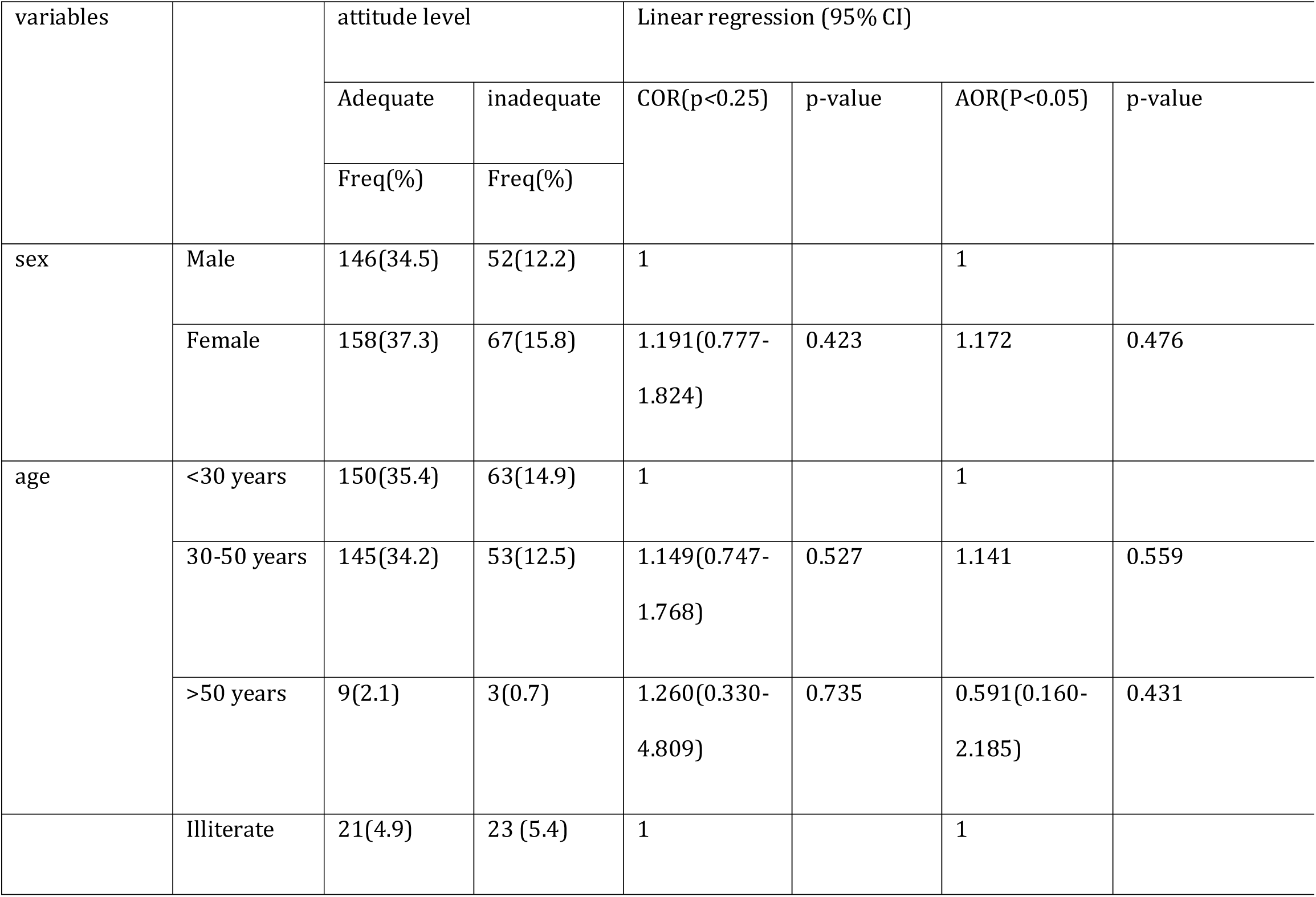
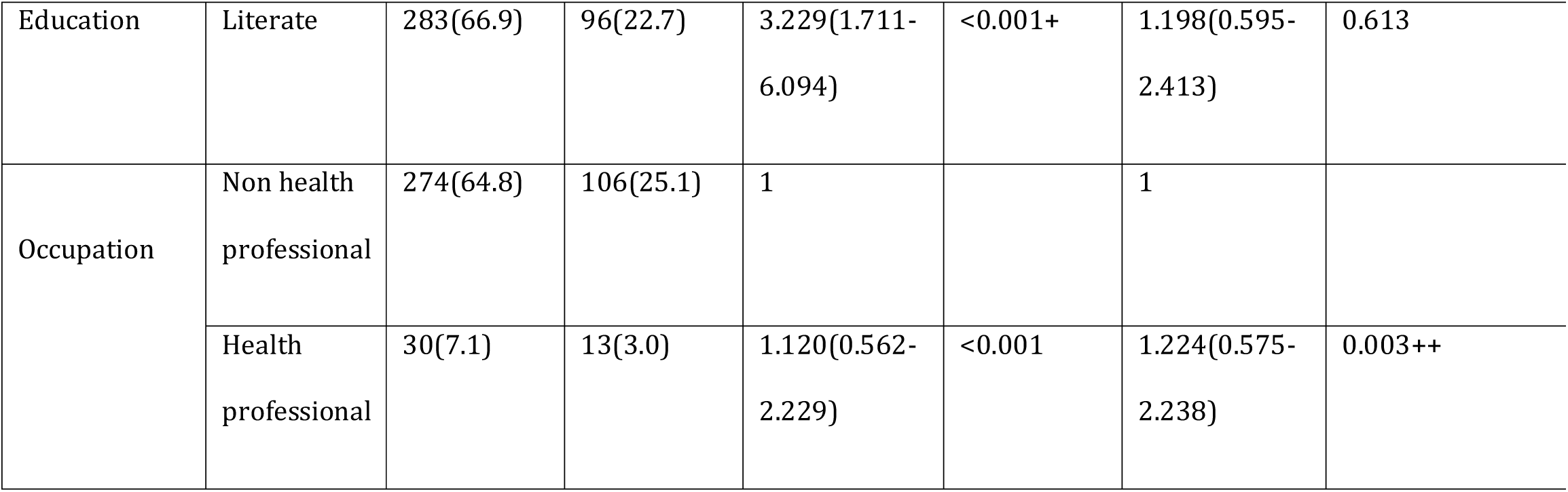
Binary and multiple logistic regression analysis of selected factors affecting attitude on FBA and obstruction.

**TABLE 7:**
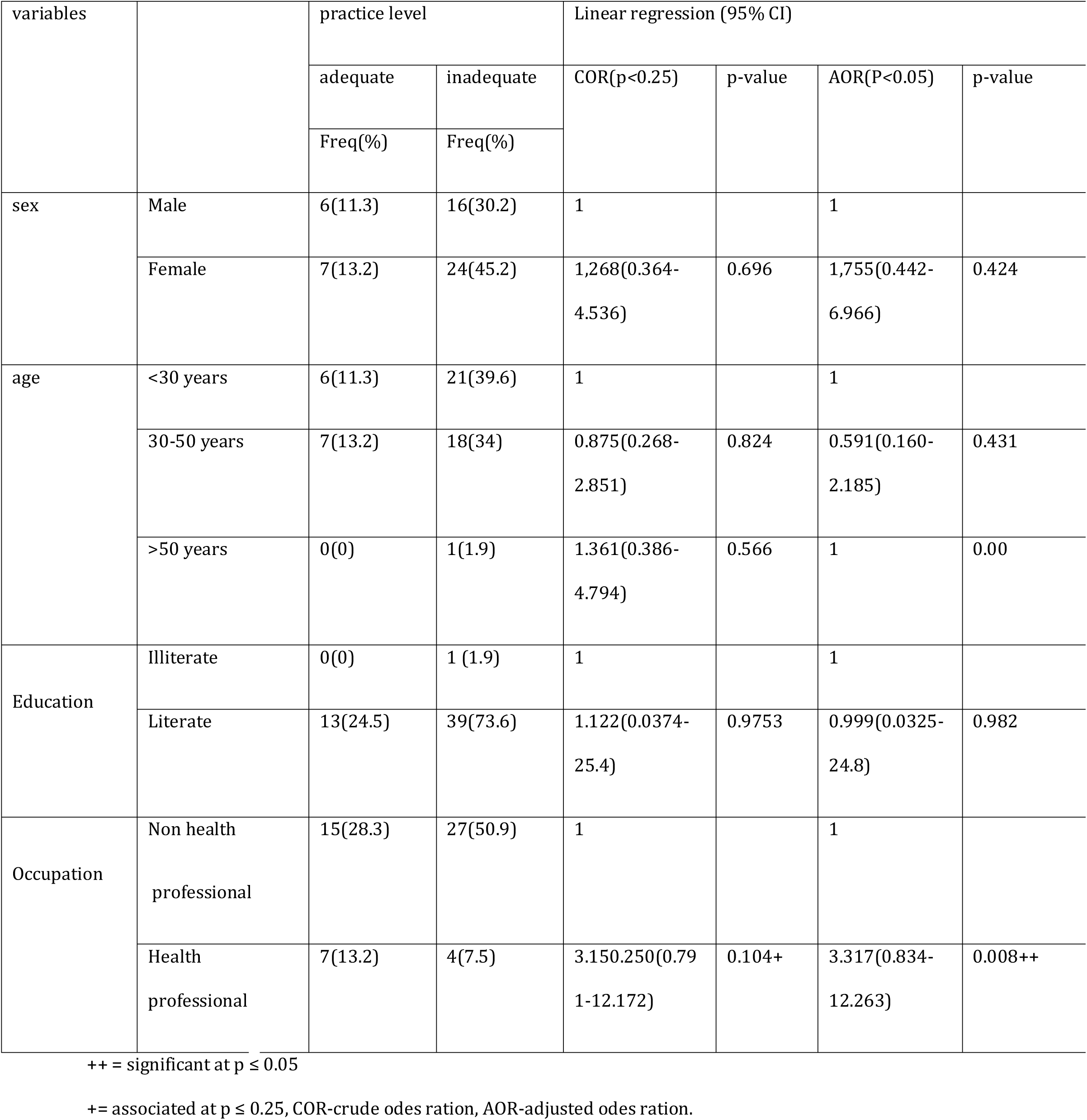
Binary and multiple logistic regression analysis of selected factors affecting practice on FBA and obstruction.

## Discussion

FBA is considered the leading cause of morbidity and mortality among young children. (15; 16) Based on the report by CDC (Center for Disease Control), it is among the leading causes of fatal home accidents in children between one and four years of age (between 2000 and 2006).

(30) Hence, assessing and increasing the awareness of parents regarding foreign body aspiration and obstruction has a vital role in decreasing the rate of morbidity and mortality associated with foreign body aspiration (10; 17; 18; 15)In this study, we measured knowledge, attitude, and practice among parents towards first aid management of foreign body aspiration and obstruction for children.

Only 51.5% of parents had adequate knowledge about foreign body aspiration and obstruction. Our results were consistent with the findings in Turkey; where 50% of the mothers who participated in the study had adequate information on foreign body aspiration. On the other hand, the study done in Al Qassim, Saudi Arabia (20) showed against our finding, where 36.9% of parents found good knowledge. In our study, a significant majority (68.8%) of parents were familiar with foreign body aspiration (FBA).

As parents’ knowledge plays an important role in lowering the incidence of FBA among children, we assessed their knowledge in our study. We found that most parents (75.7%) believed that children should not be introduced to peanuts before the age of four, and a vast majority (67.8%) recognized nuts, seeds, and plastic toys as potential aspiration hazards. These findings are consistent with several international and local studies. (2,14,20).

In our study, the majority (76.7%) of them had a positive attitude towards providing first aid for a child with foreign body aspiration and obstruction. It is slightly lower than kindergarten teachers in Addis Ababa governmental schools (17), with 95.1% having a positive attitude towards choking first aid. This difference may be due to difference in education level. In our study, 84.6% of participants agreed that foreign body aspiration needs immediate management. Similarly, a significant majority of respondents (48%) strongly agreed, and an additional 33.1% agreed, that knowledge of first-aid management for foreign body aspiration and obstruction is essential for everyone, which is comparable with kindergarten teachers (17), where 80% of the participants agreed that everyone should have basic first aid knowledge.

With regard to parents’ FBA practices, our study found that only 24.5% of the 53 parents who participated had scores above 80%. This is similar to a study conducted in Makkah (29), where only 22.4% of participants had good practices, and marginally lower than the Al Qassim study (20), which discovered that 44.7% of parents had good FBA practices and 55.3% had poor behaviors. This may be due to a low knowledgeable than those who were illiterate. (AOR:3.612, 95% CI: 1.758, 7.420, P *≤*0.05). Furthermore, a significant number of parents used risky and ineffective first aid techniques for treating patients who had aspirated foreign bodies or were obstructed, such as tapping the back of the neck, blindly sticking fingers down the victim’s throat, and offering water to drink. Based on these results, it is crucial to educate the public and make them aware of the dangers and poor behaviors that could cause this incident in order to lower the risk and incidence of FBA.

### Limitation of the study

One of the limitations is the study design being cross-sectional so, that cause and effect association cannot be studied. Second, there may be a bias related to participants’ level of understanding since the data collection tool was a self-administered questionnaire and interview for those who cannot read. Different mechanisms were tried to minimize bias by performing pre-tests before actual data collection.

### Conclusion

As there is very few study in our country, the purpose of this research was to examine the parents knowledge, attitudes, and practice toward the first aid management of foreign body aspiration and obstruction. It is hoped that the information gained from this study will assist the concerned bodies who are responsible. Since foreign body aspiration in children is an avoidable condition preventive care awareness by health care professionals in general and training and educating of the families is very important. Foreign body aspiration awareness needs to be enhanced, especially among mothers, through the use of both visual and verbal tools to reduce mortality and prevent complications. Women should be the primary target population for this educational initiative, as higher female educational attainment is associated with improved public welfare.

### Recommendations

This study identified low awareness and practice regarding foreign body aspiration (FBA) among the target population, likely attributed by high illiteracy rates. So based on the identified gaps form the study the following recommendation are made:

- Most importantly, physicians and health care workers can play a significant role in spreading awareness about FBA in the community. Sensitizing the community toward the need of awareness regarding FBA through educational campaigns involving during routine visits to the health care settings.
- Educational campaign or educational message should be carried out through television and radio broadcasts, leaflets, interviews in newspapers, community gatherings, and mobile phone messaging services to reach a wider audience.
- Creating visual aids (pictograms, short videos, demonstrations) alongside translated messages in local languages to overcome literacy barriers.
- Integrate FBA awareness into existing programs like parenting workshops, maternal health initiatives, and early childhood development programs.

## Data Availability

All relevant data are within the manuscript and its Supporting Information files.

## Acknowledgments

We thank SPHMMC for providing us to conduct this study and its ethical approval. We also thank to Supervisor, Data collectors and study participants for their immense cooperation during data collection period.

## Author Contributions

**Conceptualization:** Yohannes Hailu, Sisay Yifru

**Data curation:** Yohannes Hailu, Sisay Yifru

**Formal analysis:** Yohannes Hailu

**Investigation:** Yohannes Hailu

**Methodology:** Yohannes Hailu, Sisay Yifru

**Software:** Yohannes Hailu, Sisay Yifru

**Supervision:** Sisay Yifru

**Validation:** Yohannes Hailu, Sisay Yifru

**Visualization:** Yohannes Hailu, Sisay Yifru

**Writing – original draft:** Yohannes Hailu

**Writing – review & editing:** Yohannes Hailu, Sisay Yifru

## Notes

### Competing Interest Statement

The authors have declared no competing interest.

### Funding Statement

The author(s) received no specific funding for this work.

### Author Declarations

Ref. No. RM231926 Date: 13/6/2025 Institutional Review Board (IRB) of St. Paul's Hospital Millennium Medical College (SPHHMC) Ethical Clearance Research Title: Assessment of Knowledge, Attitude, And practice towards the first aid management of foreign body aspiration and obstruction among parents of children visited SPHMMC Addis Ababa, Ethiopia Principal Investigator: Dr:Yohannes Hailu The IRB of SPHMMC has reviewed the above mentioned research proposal and made the following decision: • Approved:- The decision is valid for 12 Approved with recommendation:- Approved on condition :- Disapproved:- months and the research should be conducted in compliance with the protocol/proposal approved by the IRB of SPHMMC. Any subsequent revision/amendment of the protocol/proposal needs approval before conduct of the research. The researcher should also submit written summaries of the research status to the IRB every 03 months. Upon the conclusion of the study, manuscripts and thesis work to the final/completed research project needs to be submitted to the IRB.

